# Prevalence and Time of Diagnosis of Endometriosis across Racial and Ethnic Groups in the US

**DOI:** 10.1101/2021.07.28.21261303

**Authors:** Rui Li, Donna A Kreher, Ashley L Gubbels, Amy R Benjamin

## Abstract

**Background:** Endometriosis is associated with substantial adverse health and psychosocial consequences. However, the prevalence of endometriosis across different racial and ethnic groups in the US has not been established and to what extent the difference in prevalence might be associated with systemic barriers to receiving the diagnosis is unknown. We aimed to estimate the prevalence of endometriosis across different racial and ethnic groups and examine the association between race/ethnicity and the time of diagnosis

**Methods:** Using cross-sectional data from the National Health and Nutrition Examination Survey 1999-2006, we estimated race/ethnicity-specific prevalence of self-reported diagnosis of endometriosis among 5,557 women aged 20-54 years. Multivariable generalized linear regression model examined the association between race/ethnicity and the age of endometriosis diagnosis, accounting for socioeconomic indicators, health insurance, lifestyle and reproductive characteristics.

**Results:** Endometriosis was prevalent in 9.0% overall, 11.1% among non-Hispanic white, 5.8% among non-Hispanic black, 2.7% among Hispanic, and 6.4% among the other racial and ethnic groups combined. The mean age of endometriosis diagnosis was 29.0 years (range 13-52). Compared with non-Hispanic white, non-Hispanic black women were diagnosed on average at 2.6 years older (95% confidence interval [CI], 0.5-4.6), Hispanic women 3.8 years older (95% CI 1.5-6.2), and women in the other group 1.0 years younger (95% CI, -4.6 to 2.5).

**Conclusions:** Prevalence of endometriosis varied across different racial and ethnic groups Among women with a diagnosis of endometriosis, black and Hispanic women were diagnosed at older ages than non-Hispanic white women. Future research is needed to determine whether such age disparity in endometriosis diagnosis persists.

## Introduction

Endometriosis is a chronic gynecological condition characterized by the presence of tissue with the histologic appearance of endometrial glands and stroma outside of the endometrial cavity, which often causes dysmenorrhea, dyspareunia, chronic pelvic pain, and is often associated with infertility (Howard, 2009). It is suggested to be prevalent in about 10% of women of reproductive age (Shafrir et al., 2018). With $12,118 direct cost and $15,737 indirect cost per patient per year, the economic toll of endometriosis is highest in the United States (US) (Ahmed M. Soliman, Yang, Du, Kelley, & Winkel, 2016). A few studies have estimated the prevalence of endometriosis among reproductive-aged women in the US, although these estimates may not be an accurate reflection of the nationwide prevalence due to study design limitations such as endometriosis assessment method and self-selection into online survey (Bernuit et al., 2011; Fuldeore & Soliman, 2017; Kjerulff, Erickson, & Langenberg, 1996). Importantly, the prevalence of endometriosis within various racial and ethnic groups in the US is still unknown despite the conclusion from a recent systematic review that Black and Hispanic women were less likely to be diagnosed with endometriosis than white women (Bougie, Yap, Sikora, Flaxman, & Singh, 2019).

Disparity in endometriosis diagnosis may contribute to the commonly believed and observed lower prevalence of endometriosis among nonwhite women. Such racial disparity could be due in part to socioeconomic factors such as access to health care, as the diagnosis of endometriosis is usually preceded by multiple clinical visits sometimes between numerous providers, and requires laparoscopic confirmation. Extant literature suggests that the delay from symptom onset to the diagnosis of endometriosis ranges from 4-11 years (Agarwal et al., 2019; Nnoaham et al., 2011; A. M. Soliman, Fuldeore, & Snabes, 2017), with significant delay even in countries with universal health care (Hudelist et al., 2012; Staal, van der Zanden, & Nap, 2016). In the US where the health care systems are largely segregated and the health care resources are unevenly distributed across different socioeconomic strata (Fiscella & Williams, 2004; Ko, Needleman Derose, Laugesen, & Ponce, 2014; Smith, 2005), women of different racial and ethnic backgrounds may face different levels of challenges on their path to the diagnosis of endometriosis. However, there has been little research in the experiences of the diagnosis of endometriosis in general, with even less research focusing on the experiences of racial and ethnic minorities

In addition to socioeconomic factors, diagnostic biases may also play a role in explaining for the racial disparity in the diagnosis. Endometriosis has historically been depicted as a disease of white and affluent women who tend to postpone childbearing and is believed to be rare among nonwhite women (Blinick & Merendino, 1951; Meigs, 1938; Scott & Te Linde, 1950). This notion originated from patient observations among clinics that mainly served patients with private insurance and/or white women (Blinick & Merendino, 1951; Gibson & Jung, 2005), and is bolstered by the prevalence data that suggest a lower prevalence of endometriosis among nonwhite women (Bougie, Yap, et al., 2019; Fuldeore & Soliman, 2017; Mowers et al., 2016; Nnoaham et al., 2011). However, these prevalence estimates only provide a snapshot view of endometriosis presence among populations and fail to take into account the diagnostic process. The prevailing notion of white predominance of the disease may perpetuate the diagnostic bias in medical professionals, which is evidenced by the observation that 40% of African-American women with a laparoscopic evidence of endometriosis were previously misdiagnosed with pelvic inflammatory disease (Chatman, 1976). To date, to what extent the seemingly lower prevalence of endometriosis among nonwhite women is attributed to potential diagnostic biases remains unknown, impeding the goal of early diagnosis and quality care of endometriosis among minority female populations.

To gain a better understanding of both the epidemiology and diagnosis of endometriosis across different racial and ethnic groups in the US, we estimated the race and ethnicity-specific prevalence of endometriosis among US women of reproductive age and further examined the association between race/ethnicity and the age of endometriosis diagnosis, using data from the National Health and Nutrition Examination Survey (NHANES). We examined this association by conditioning on socioeconomic, lifestyle, and reproductive characteristics, so that the difference found across racial and ethnic groups in the age of diagnosis could potentially reflect racially and/or ethnically-related diagnostic biases for endometriosis.

## Methods

### Data Source

We used publicly available data from the NHANES, a cross-sectional study that uses a complex, multistage, probability sampling design to survey a nationally representative sample of about 10,000 noninstitutionalized US residents in 2-year cycles (Centers for Disease Control and Prevention (CDC). National Center for Health Statistics (NCHS).). NHANES interview data include demographic, socioeconomic, dietary, and health-related questions. Participants are interviewed by trained technicians using standardized questionnaires and methods, which ensures rigorous interview standards and high data quality. We used combined data from the 1999-2000, 2001-2002, 2003-2004 and 2005-2006 NHANES cycles because questions regarding endometriosis were asked during these cycles. Response rates of NHANES 1999-2006 ranged

between 76–80%. Since we used publicly available deidentifed data, the university Institutional Review Board exempted the current study from review (STUDY00005239).

### Study Population

For estimating the prevalence of endometriosis, our study population consisted of women aged between 20-54 years who participated in NHANES 1999-2006 and completed the reproductive health questionnaire in which they gave answers to the endometriosis questions. From 1999-2006, NHANES oversampled non-Hispanic black and Mexican-American individuals. Over 85% of the Hispanic participants were Mexican-American individuals. For examining the association between race/ethnicity and the age of endometriosis diagnosis, the study population were women who reported ever receiving a diagnosis of endometriosis and gave information regarding the age of diagnosis.

### Measurement

Endometriosis was ascertained from the reproductive health questionnaire in which women were asked “Has a doctor or other health professional ever told you that you had endometriosis?”. Women who answered *yes* to this question were further asked “How old were you when you were first told you had endometriosis?” with the age of diagnosis measured in years.

The race/ethnicity variable was derived by combining responses to questions on race and Hispanic origin. Respondents who self-identified as “Mexican American” or “Other Hispanic” were coded as being Hispanic. All other non-Hispanic participants were categorized based on their self-reported races: non-Hispanic white, non-Hispanic black, and other non-Hispanic race including non-Hispanic multiracial.

Covariates for examining the association between race/ethnicity and age of endometriosis diagnosis included socioeconomic indicators which may affect both the incidence of endometriosis and the access to health care, and lifestyle and reproductive characteristics which primarily influence the risk of endometriosis incidence. Many of these variables potentially lie on the causal pathway between race/ethnicity and age of endometriosis diagnosis; by accounting for these variables we were able to disentangle the effect of race/ethnicity on the time of diagnosis that is independent of socioeconomic status (including access to health care), lifestyles, and reproductive factors.

Socioeconomic indicators included education attainment (less than high school, high school diploma, and more than high school), family income-to-poverty ratio which ranged from 0-5 with 0 indicating no income and 5 indicating ≥ 5 times the federal poverty level, and type of health insurance (private insurance, public insurance including Medicaid/CHIP and other government insurance, and uninsured). Lifestyle variables included smoking history and body mass index (BMI). For smoking history, we created a binary variable indicating whether women started regular smoking before the age of 18 years. We chose this cutoff because the majority of smokers started smoking before 18 (U.S. Department of Health and Human Services, 2012) and also to minimize the reversal effect of a diagnosis of endometriosis on smoking. For BMI, we used women’s self-reported current height and maximum lifetime weight (not including weight during pregnancy) to construct a variable of maximum lifetime BMI (kg/m^2^), which was further categorized into <25 kg/m^2^, 25–29.9 kg/m^2^, ≥ 30 kg/m^2^. Reproductive characteristics included age of menarche (years), parity (parous or not), and history of birth control pills use. For birth control pills use, we calculated the duration (years) of use prior to the diagnosis of endometriosis in the following way: for women who did not report taking any birth control pills or initiated birth control pills after the diagnosis of endometriosis, their duration of use was 0; for women who initiated birth control pills prior to the diagnosis of endometriosis, their duration of use was calculated as either the total years of birth control pills use if they stopped prior to the diagnosis of endometriosis, or the age of endometriosis diagnosis minus the age of birth control pills initiation if they continued the use after the diagnosis of endometriosis.

### Statistical Analyses

We calculated cycle-specific prevalence of endometriosis, accounting for the complex sample design by applying cycle-specific sampling weights and sample design variables. We then calculated the pooled overall prevalence as well as the prevalence for different racial and ethnic groups from 1999-2006 using calibrated sample weights and sample design variables (Centers for Disease Control and Prevention (CDC). National Center for Health Statistics (NCHS).)

We compared the mean age of endometriosis diagnosis by racial and ethnic groups, socioeconomic, lifestyle, and reproductive characteristics using the classical analysis of variance (ANOVA) when the assumption of homogenous variance was met or Welch’s ANOVA when this assumption was violated (Welch, 1951). To facilitate comparison across categories, we categorized family income-to-poverty ratio into <1.00, 1.00-1.99, 2.00-4.99, and 5.00-highest; age of menarche into 8-11, 12-13, and 14-17 years; and years of birth control pills use into none, < 5 years, and ≥ 5 years, and reported the mean age of diagnosis for each category.

To examine the association between race/ethnicity and age of endometriosis diagnosis, we conducted a generalized multivariable linear regression model, adjusting for socioeconomic, lifestyle, and reproductive factors, so that the association observed between race/ethnicity and age of endometriosis diagnosis could potentially reflect the difference due to the diagnostic bias. Data management and statistical analyses were conducted in SAS 9.4 (SAS Inc., Cary, NC, USA).

## Results

The flowchart for sample selection is presented in Figure 1. Out of 41,474 NHANES participants 1999–2006, 6,508 were women aged 20-54 years and 95.4% of them completed the reproductive health questionnaire among whom 89.5% had information regarding whether they had been diagnosed of endometriosis. From 1999–2006, the prevalence of endometriosis ranged between 8.1% and 9.4% (Table 1), with a pooled prevalence of 9.0%. The prevalence was highest among non-Hispanic white women (11.1%) and lowest among Hispanic women (2.7%).

**Figure 1.**
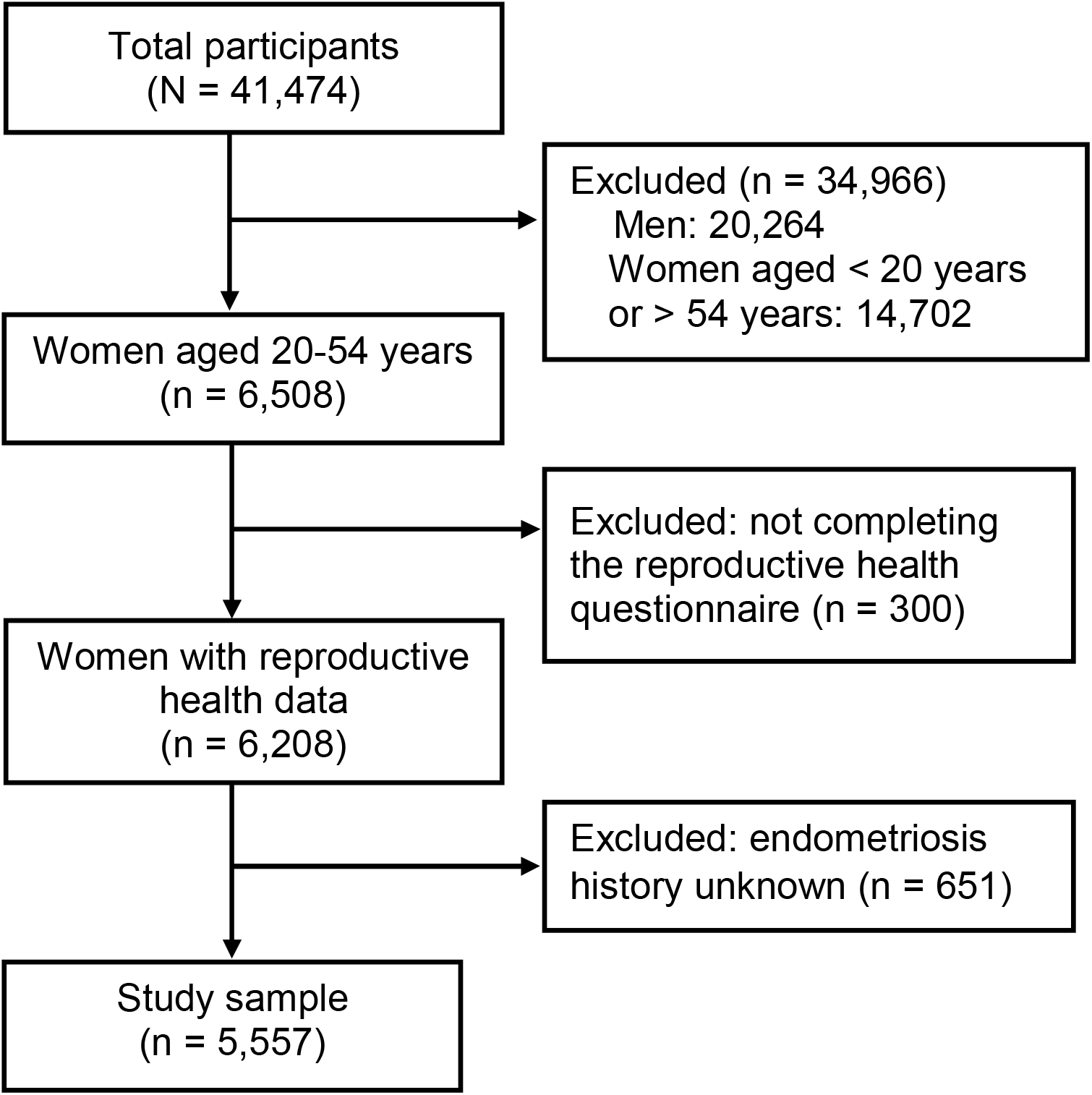
Flowchart for sample selection

**Table 1.** Prevalence of Self-reported Diagnosis of Endometriosis among US Women Aged 20-54 Years, Results from NHANES 1999-2006 (Unweighted n = 5,557)

|  | **Unweighted** | **Unweighted** | **Weighted** |  |
| --- | --- | --- | --- | --- |
|  | **total** | **endometriosis** | **prevalence** | **95% CI** |
|  | **n** | **n** | **(%)** |  |
| 1999-2000 | 1,325 | 84 | 9.4 | 6.0-12.9 |
| 2001-2002 | 1,486 | 108 | 9.2 | 7.0-11.4 |
| 2003-2004 | 1,275 | 84 | 8.1 | 6.4-9.9 |
| 2005-2006 | 1,471 | 104 | 9.3 | 6.8-11.8 |
| Pooled 1999-2006 | 5,557 | 380 | 9.0 | 7.9-10.2 |
| *Non-Hispanic White* | 2,568 | 261 | 11.1 | 9.5-12.7 |
| *Non-Hispanic Black* | 1,151 | 67 | 5.8 | 4.5-7.1 |
| *Hispanic* | 1,585 | 37 | 2.7 | 1.4-4.0 |
| *Others* | 253 | 15 | 6.4 | 3.1-9.7 |

CI = confidence interval; NHANES = National Health and Nutrition Examination Survey.

Among the 380 women who reported having a diagnosis of endometriosis, age of diagnosis was available for 375 of them. As shown in Table 2, the mean age of diagnosis was 29 years. A later diagnosis of endometriosis was seen among black and Hispanic women, women with a high school degree, women without health insurance, women with family income below the poverty line, women who did not smoke and who had a higher BMI, women who had a later age of menarche, who were parous and who reported the longest duration of birth control pills use prior to the diagnosis of endometriosis.

**Table 2.** Age of Endometriosis Diagnosis by Race/Ethnicity, Socioeconomic, Health Insurance, Lifestyle and Reproductive Characteristics (n = 375)

| Characteristic | N | Age of diagnosis:<br>Mean (±SD) | p value<sup>a</sup> |
| --- | --- | --- | --- |
| Mean age of endometriosis diagnosis | 375 | 29.0 (±8.1) | |
| Race and ethnicity |  |  | 0.126 |
| Non-Hispanic White | 259 | 28.5 (±7.9) | |
| Non-Hispanic Black | 64 | 30.1 (±8.7) | |
| Hispanic | 37 | 31.2 (±8.4) | |
| Others | 15 | 27.3 (±6.1) | |
| Education level |  |  | 0.281 |
| Less than high school | 47 | 29.1 (±8.8) | |
| High school diploma, GED | 100 | 30.0 (±7.5) | |
| More than high school | 228 | 28.5 (±8.1) | |
| Type of insurance |  |  | 0.558 |
| Private | 269 | 28.7 (±7.8) | |
| Public | 51 | 29.1 (±9.2) | |
| Uninsured | 55 | 30.0 (±8.2) | |
| Income-to-poverty ratio |  |  | 0.159 |
| <1.00 | 49 | 30.1 (±8.2) | |
| 1.00-1.99 | 61 | 29.1 (±8.9) | |
| 2.00-4.99 | 159 | 27.9 (±7.7) | |
| 5.00-highest | 93 | 29.9 ($\pm 8.0$) | |
| Smoked regularly before age 18 |  |  | 0.314 |
| Yes | 90 | 28.2 ($\pm 7.3$) | |
| No | 285 | 29.2 ($\pm 8.3$) | |
| Highest BMI not including pregnancy: kg/m<sup>2</sup> |  |  | 0.475 |
| < 25 | 105 | 28.2 ($\pm 7.6$) | |
| 25–29.9 | 104 | 29.0 ($\pm 8.1$) | |
| $\geq 30$ | 160 | 29.5 ($\pm 8.3$) | |
| Age of menarche (years) |  |  | 0.091 |
| 8-11 | 98 | 28.0 ($\pm 8.3$) | |
| 12-13 | 198 | 28.7 ($\pm 8.0$) | |
| 14-17 | 74 | 30.6 ($\pm 7.6$) | |
| Parous prior to the diagnosis of endometriosis |  |  | <.0001 |
| Yes | 191 | 32.6 ($\pm 7.2$) | |
| No | 184 | 25.2 ($\pm 7.2$) | |
| Years of birth control pills use prior to the diagnosis of endometriosis |  |  | <.0001<sup>b</sup> |
| None | 128 | 26.4 ($\pm 8.2$) | |
| < 5 years | 114 | 28.1 ($\pm 8.1$) | |
| $\geq 5$ years | 131 | 32.2 ($\pm 6.8$) | |

GED = General Educational Diploma; BMI = body mass index; SE = standard errors.
<sup>a</sup> Means were compared using the analysis of variance (ANOVA).
<sup>b</sup> Welch's ANOVA was used due to the violation of homogeneity of variance.

Adjusting for socioeconomic, lifestyle, and reproductive factors, there was a statistically significant difference in the age of endometriosis diagnosis across different racial and ethnic groups (Table 3). Compared with non-Hispanic white women, non-Hispanic black women were on average diagnosed of endometriosis 2.62 years later and Hispanic women were diagnosed 3.85 years later. Women identifying as “Other” were diagnosed slightly earlier but the difference was not significant. In this adjusted model, income-to-poverty ratio, being parous, and years on birth control pills were also significant predictors of age of endometriosis diagnosis. Particularly, parous women were diagnosed of endometriosis 7 years later than nulliparous women.

**Table 3.** Multivariable Generalized Linear Regression Modeling of the Association between Race/Ethnicity and Age of Endometriosis Diagnosis (n = 351)

| Predictors | Beta coefficient | 95% CI | p value |
| --- | --- | --- | --- |
| Race/Ethnicity |  |  | .002 |
| Non-Hispanic white | Ref | Ref |  |
| Non-Hispanic black | 2.62 | 0.64-4.60 |  |
| Hispanic | 3.85 | 1.48-6.21 |  |
| Other groups combined | -1.04 | -4.57 to 2.50 |  |
| Education level |  |  | .669 |
| Less than high school | Ref | Ref |  |
| High school diploma, GED | 1.05 | -1.41 to 3.50 |  |
| More than high school | 0.48 | -1.82 to 2.78 |  |
| Family income to poverty ratio | 0.59 | 0.04-1.13 | .035 |
| Type of insurance |  |  | .188 |
| Private | Ref | Ref |  |
| Public | 1.17 | -1.20 to 3.54 |  |
| Uninsured | 2.08 | -0.20 to 4.36 |  |
| Smoked regularly before age 18 | -1.16 | -2.84 to 0.53 | .178 |
| Highest BMI not including pregnancy: kg/m<sup>2</sup> |  |  | .715 |
| < 25 | Ref | Ref |  |
| 25–29.9 | 0.74 | -1.08 to 2.56 |  |
| ≥ 30 | 0.51 | -1.22 to 2.24 |  |
| Age of menarche (years) | 0.31 | -0.09 to 0.72 | .125 |
| Parous: yes vs no | 7.01 | 5.50-8.52 | <.0001 |
| Years of birth control pills use prior to the<br>diagnosis of endometriosis | 0.49 | 0.35-0.62 | <.0001 |

GED = General Educational Diploma; BMI = body mass index; CI = confidence interval.

## Discussion

In this large, nationally representative sample of US women of reproductive age surveyed between 1999-2006, the prevalence of self-reported diagnosis of endometriosis was 9.0%, consistent with the commonly quoted 10% prevalence. The prevalence was highest among non-Hispanic white women and lowest among Hispanic women. Accounting for socioeconomic, lifestyle, and reproductive characteristics, non-Hispanic women were diagnosed with endometriosis at 2.6 years older, and Hispanic women at 3.8 years older, compared with non-Hispanic white women.

This study is among the few to estimate the prevalence of endometriosis in the US. Our national prevalence estimate of 9% is higher than the 12-month prevalence of 6.9% reported by Kjenilff et al using data from the National Health Interview Survey 1984-1992 (Kjerulff et al., 1996). Two online surveys conducted in 2009 and 2012 among the US women aged 18-49 reported the prevalence of endometriosis to be 6.6%–10.8% (Bernuit et al., 2011) and 6.1% (Fuldeore & Soliman, 2017), respectively. However, the selected participants only represented 74% of the target US population in the first study and neither study addressed selection bias due to the nature of online surveys. Our prevalence was estimated from a nationally representative sample which is less subject to selection bias.

This study is the first to describe the national prevalence of endometriosis by different racial and ethnic groups in the US. Non-Hispanic white women had twice the prevalence compared to non-Hispanic black women. The prevalence estimate among Hispanic women in our study (2.7%) was lower than the reported 4% among Puerto Rican women (Flores et al., 2008). This may be due to different sampling methods (complex, multistage, probability sampling in ours vs convenient sampling in the latter) and different population characteristics (predominantly Mexican American women in ours vs Puerto Rican women in the latter).

Beyond confirming a higher prevalence of endometriosis among non-Hispanic white women compared with non-Hispanic Black and Hispanic women, our study makes a significant contribution by revealing, for the first time, the gaps in the age of endometriosis diagnosis across different racial and ethnic groups, potentially reflecting the racially and ethnically-related diagnostic biases of endometriosis. Different racial and ethnic groups may have different risks of developing endometriosis as suggested by a prospective cohort study (Missmer et al., 2004), and such difference could be due to genetic, lifestyle, and reproductive factors (Shafrir et al., 2018). The association between socioeconomic factors and endometriosis may be more complex, as these factors may influence both endometriosis incidence and access to health care. An earlier diagnosis of endometriosis among those with health care access does not mean an earlier onset of endometriosis. Our findings, after conditioning on socioeconomic, lifestyle, and reproductive factors, demonstrated a 2.6 and 3.8 years’ difference in the age of endometriosis diagnosis for black and Hispanic women relative to white women, respectively. This difference could be due to a true late onset of endometriosis among black and Hispanic women, different symptoms or clinical presentations of endometriosis among these women, or potentially biases among health care providers making the diagnosis (Bougie, Healey, & Singh, 2019). Since we controlled for the most established risk factors of endometriosis (i.e., BMI, age of menarche, parity, birth control pills use), as well as socioeconomic factors (including health insurance as an indicator of health care access), we believe that the observed gap in the age of endometriosis diagnosis partly reflects race/ethnicity-associated diagnostic biases.

Endometriosis in women is associated with substantial negative health, developmental (e.g., pursuing education and career choices, starting a family), occupational, and relational impact across the lifespan (Missmer et al., 2021), but many women experience significant delay in receiving the diagnosis. According to a recent online survey of women with endometriosis, more than half waited 6 or more years for the diagnosis of endometriosis while 54.3% experienced endometriosis-related pain daily (Lamvu, Antunez-Flores, Orady, & Schneider, 2020). The delay in the diagnosis has been reported to occur at both the individual patient level and the medical community level (Ballard, Lowton, & Wright, 2006). For example, it is very common for women to normalize and accommodate their menstrual pain which is usually the first symptom of endometriosis. At the medical level, there is a significant delay in referral from primary to secondary care which may be associated with pain normalized by family doctors. Intermittent hormonal suppression of symptoms as well as the use of nondiscriminatory diagnostic tools further contribute to the delay (Ballard et al., 2006). Relating to the discrepancy in the age of endometriosis diagnosis across different racial and ethnic groups found in our study, there could be a greater degree of normalization of menstrual symptoms among black and Hispanic women themselves as well as medical professionals in treating these patients. Additionally, due to the commonly held belief that endometriosis is less common among nonwhite women, providers may be less likely to associate symptoms in black and Hispanic women with endometriosis, and therefore, less likely to make a timely and proper referral.

Limitations for the present study must be noted. First, we were not able to conclude definitely whether the observed differences in the age of endometriosis diagnosis were due to different disease/symptom onset or delay in diagnosis, although we controlled for common risk factors of endometriosis. Future research is needed to examine the duration from symptom onset to a confirmatory clinical diagnosis of endometriosis across various racial and ethnic groups to delineate the diagnostic disparity that could be avoided. Second, we were not able to ascertain the diagnostic method (e.g., histologically proven, surgical, clinical suspicion) and presenting symptoms (e.g., with or without pain, with or without infertility) of endometriosis. It is possible that black and Hispanic women were diagnosed with endometriosis by chance at older ages.

Future research should identify and focus on the clinical meaningful phenotypes of endometriosis that are relevant to women of different racial and ethnic background. Third, for hormonal contraceptives, we only controlled for birth control pills use which was the only type of contraceptive that we were able to ascertain the length of use prior to the diagnosis of endometriosis, which may cause residual confounding. Fourth, we analyzed the NHANES 1999-2006 data. Over the years there has been an increasing awareness of endometriosis in the medical field and the society. More research is needed to determine whether the gap in the age of endometriosis diagnosis persists across different racial and ethnic groups, and if so, whether factors contributing to this diagnostic disparity change over time.

### Implications for Practice and/or Policy

Clinicians should be aware that the historically held belief that endometriosis is rare among nonwhite women may be misleading and fueling an inherent race/ethnicity-related bias in making the diagnosis, thereby creating a diagnostic disparity across different racial and ethnic groups and exacerbating the delay in receiving appropriate care for black and Hispanic women. Primary care physicians should pay attention to the menstrual symptoms of black and Hispanic women, who may themselves normalize symptoms suggestive of underlying endometriosis, and make a timely referral for suspicious endometriosis. It is important to communicate to black and Hispanic patients that endometriosis can happen in all racial and ethnic groups, and severe menstrual and/or pelvic pain symptoms should not be normalized. Clinicians should also consider the possibility that women of different racial and ethnic background could have different symptoms and clinical presentations of endometriosis, and therefore, atypical symptoms should not rule out the diagnosis but warrant close examination. Finally, it is imperative to dispel the belief in the medical community that endometriosis is a white disease so that implicit racial biases in the diagnosis of endometriosis could be eliminated and treatment decisions are not influenced by such biases (Bougie, Healey, et al., 2019).

## Conclusions

Self-reported endometriosis was prevalent in 9% of reproductive-aged US women between 1999 and 2006. Black and Hispanic women had lower prevalence of endometriosis, and were diagnosed with endometriosis at later ages, compared with their white counterparts. This later diagnosis was observed after accounting for factors associated with the risk of and access to care for endometriosis, which potentially reflects the diagnostic biases related to race and ethnicity.

## Data Availability

We used publicly available data from the NHANES which were downloaded from the website (https://wwwn.cdc.gov/nchs/nhanes/Default.aspx).

